# Dysbiosis and structural disruption of the respiratory microbiota in COVID-19 patients with severe and fatal outcomes

**DOI:** 10.1101/2021.05.19.21257485

**Authors:** Alejandra Hernández-Terán, Fidencio Mejía-Nepomuceno, María Teresa Herrera, Omar Barreto, Emma García, Manuel Castillejos, Celia Boukadida, Margarita Matias-Florentino, Alma Rincón-Rubio, Santiago Avila-Rios, Mario Mújica-Sánchez, Ricardo Serna-Muñoz, Eduardo Becerril-Vargas, Cristobal Guadarrama-Pérez, Víctor Hugo Ahumada-Topete, Sebastián Rodríguez, José Arturo Martínez-Orozco, Jorge Salas-Hernández, Rogelio Pérez-Padilla, Joel Armando Vázquez-Pérez

**Author notes:** **Corresponding author**: Joel Armando Vázquez-Pérez.

## Abstract

COVID-19 outbreak has caused over 3 million deaths worldwide. Understanding disease pathology and the factors that drive severe and fatal clinical outcomes is of special relevance. Studying the role of the respiratory microbiota in COVID-19 is particularly important since it’s known that the respiratory microbiota interacts with the host immune system, contributing to clinical outcomes in chronic and acute respiratory diseases. Here, we characterized the microbiota in the respiratory tract of patients with mild, severe, or fatal COVID-19, and compared with healthy controls and patients with non-COVID-19-pneumonia. We comparatively studied the microbial composition, diversity, and microbiota structure across study groups and correlated the results with clinical data. We found differences in diversity and abundance of bacteria between groups, higher levels of dysbiosis in the respiratory microbiota of COVID-19 patients (regardless of severity level), differences in diversity structure among mild, severe, and fatal COVID-19, and the presence of specific bacteria that correlated with clinical variables associated with increased mortality risk. Our data suggest that host-related and environmental factors could be affecting the respiratory microbiota before SARS-CoV-2 infection, potentially compromising the immunological response of the host against disease and promoting secondary bacterial infections. For instance, the high levels of dysbiosis coupled with low microbial structural complexity in the respiratory microbiota of COVID-19 patients, possibly resulted from antibiotic uptake and comorbidities, could have consequences for the host and microbial community level. Altogether, our findings identify the respiratory microbiota as a potential factor associated with COVID-19 severity.

## Introduction

The Coronavirus Disease 2019 (COVID-19) outbreak, declared a pandemic by the World Health Organization on the 11th of March 2020, is caused by the Severe Acute Respiratory Syndrome Coronavirus 2 (SARS-CoV-2). As of May 2021, SARS-CoV-2 has infected more than 150 million people and caused over 3 million deaths worldwide^1^. COVID-19 shows a wide spectrum of clinical manifestations ranging from asymptomatic infection and mild respiratory symptoms to severe pneumonia and death, ^2,3^ which has been related to demographic factors and comorbidities. To date, it has been shown that the aberrant immune response against SARS-CoV-2 antigens is critically involved in severe clinical outcomes and other secondary inflammatory conditions that remain after COVID-19 ^3,4^.

Studying the role of the human microbiota in COVID-19 is particularly relevant since it’
ss known that the respiratory microbiota interacts with the host immune system, contributing to clinical outcomes in chronic and acute respiratory diseases ^5^. The respiratory microbiota has a central role in shaping pulmonary immunity by boosting innate and adaptive immune responses. Suggesting that host immunity is regulated by interactions with the bacterial communities in the respiratory tract.

Some studies suggest that the interactions between microorganisms and the host immune system are species-specific, denoting that even minor variations in the diversity and composition of the microbiota could have significant consequences on host’shealth ^6^. For COVID-19, severe to fatal clinical outcomes are often associated with the presence of comorbidities that are known to display altered (dysbiotic) microbiota ^7^ (e.g., diabetes type II, obesity, age, and heart disease). Furthermore, in a wide range of microbiome-associate diseases (MADs), dysbiosis is a common feature that can impact disease progression ^8,9^.

Nonetheless, few studies characterizing the respiratory microbiota in COVID-19 and the presence of dysbiosis are available to date ^10–14^.

To gain insight into the association between respiratory microbiota and COVID-19 severity; we characterized the microbiota in the respiratory tract of patients with mild, severe, or fatal COVID-19, and compared with healthy controls and patients with non-COVID-19-pneumonia. We comparatively studied the microbial composition, diversity, and microbiota structure across study groups and correlated the results with clinical data. These analyses let us detect 1) differences in abundance of bacteria between groups, 2) higher levels of dysbiosis in the respiratory microbiota of COVID-19 patients, 3) differences in diversity structure among mild, severe, and fatal COVID-19 microbiota, and 4) the presence of specific bacteria that correlated with clinical variables associated with increased mortality risk. In summary, our results demonstrate an increasing dysbiosis of the respiratory tract microbiota in COVID-19 patients coupled with a continuous loss of microbial complexity structure from mild to fatal COVID-19 that could potentially alter clinical outcomes. Altogether, our findings identify the respiratory microbiota as a potential factor associated with COVID-19 severity.

## Results

### Study participants

Since our sample set consists of upper and lower respiratory tract samples, we kept only upper respiratory samples for the main diversity and statistical analyses. Overall, a total of 95 samples were analyzed (mild COVID-19 = 37, severe COVID-19 = 27, fatal COVID-19 = 19, healthy control = 7, and non-COVID-19-pneumonia = 5).

Demographic data, health-related characteristics, and symptomatology are described in Table 1. Overall, 52 patients were male (54.7%) with a median age of 45 years old (IQR: 21). Regarding health conditions, 58.2% of the participants presented at least one comorbidity, being DM2 (17%), hypertension (17%), smoking (17%), and obesity (35%) the most widely represented in the cohort. The median days of symptom onset were seven, and 52.6% of the individuals received antibiotic treatment before hospitalization. Furthermore, we found important associations between some health/demographic characteristics and severity. For instance, patients with fatal COVID-19 were predominantly male (73.6%, *p* = 0.01), significantly older (median= 58, *p* = 6.57e-07), with higher BMI (median= 30.4, *p* = 0.05), and most of them received previous antibiotic treatment (78.9%, *p* = 0.002) compared with patients with severe and mild COVID-19. Also, a higher number of days after symptoms onset was found in the non-COVID-19-pneumonia group (median= 10, *p* = 0.01).

**Table 1.** Demographic data of the cohort. Only upper respiratory samples (OPS and NPS) are included in the information. Abbreviations: BMI= body mass index, DM2= Diabetes Mellitus Type 2. *Respiratory diseases: either asthma, COPD, or ILD. P values denote statistical significant differences given by Wilcoxon rank-sum test (* < 0.05, ** < 0.005, *** < 0.0005).

|  | All (N= 95) | Healthy control (N=7) | COVID-19 Mild (N=37) | COVID-19 Severe (N=27) | COVID-19 Fatal (N=19) | Non-COVID-19-pneumonia (N=5) | *p* value |
| --- | --- | --- | --- | --- | --- | --- | --- |
| Age (years), median (IQR) | 45 (21) | 35 (18) | 37 (18) | 47(21) | 58(16.5) | 49(13) | 6.75e-07 *** |
| Gender |  |  |  |  |  |  |  |
| Female, n (%) | 43 (45.2%) | 5 (71.4%) | 18 (48.6%) | 12 (44%) | 5 (26.3%) | 3 (60%) | 0.016 * |
| Male, n (%) | 52 (54.7%) | 2 (28.5%) | 19 (51.3%) | 15 (55.5%) | 14 (73.6%) | 2 (40%) |  |
| Smoking, n (%) | 15 (17%) | 0 | 0 | 8 (29.6%) | 7 (36.8%) | 0 | ns |
| NA | 8 | 1 | 4 | 0 | 1 | 2 |  |
| BMI (kg/m2), median (IQR) | 27.6(6.9) | 26.2(2.6) | 27.34(7.9) | 28.9(4.6) | 30.4 (9.8) | 24.9(2.3) | 0.05 * |
| Obesity |  |  |  |  |  |  |  |
| Not obese, n(%) | 48 (50.5%) | 5 (71.4%) | 16 (43.2%) | 16 (59.2%) | 8 (42.1%) | 3 (60%) | ns |
| Class I, n (%) | 16 (61.5%) | 0 | 9 (81.8%) | 7 (77.7%) | 2 (25%) | 0 |  |
| Class II, n(%) | 7 (29.9%) | 0 | 1 (9%) | 2 (22%) | 4 (50%) | 0 |  |
| Class III, n(%) | 3 (11.5%) | 0 | 1 (9%) | 0 | 2 (25%) | 0 |  |
| NA | 21 | 2 | 12 | 2 | 3 | 2 |  |
| Comorbidities |  |  |  |  |  |  |  |
| DM2, n (%) | 15 (17%) | 0 | 3 (8%) | 7 (70.3%) | 5 (26.3%) | 0 | ns |
| Hypertension, n (%) | 15 (17%) | 0 | 3 (8%) | 4 (14.8%) | 8 (42.1%) | 0 | ns |
| Respiratory disease*, n (%) | 4 (4.5%) | 0 | 3 (8%) | 1 (3.7%) | 1 (5.2%) | 0 | ns |
| NA | 36 | 2 | 17 | 1 | 8 | 8 |  |
| Days after symptoms onset, n (IQR) | 7 (6) | NA | 5 (5) | 6.5 (4.7) | 5 (5) | 10 (3) | 0.01* |
| Antibiotic treatment, n (%) | 50 (52.6%) | 1 (14.2%) | 11 (29.7%) | 21 (77.7%) | 15 (78.9%) | 2 (40%) | 0.002** |
| Symptoms |  |  |  |  |  |  |  |
| Cough, n (%) | 50 (52%) | 2 (28.5%) | 11 (29.7%) | 20 (74%) | 15 (78.9%) | 2 (40%) | ns |
| Fever, n (%) | 48 (50.5%) | 2 (28.5%) | 10 (27%) | 19 (70.3%) | 15 (78.9%) | 2 (40%) | ns |
| Dyspnea, n (%) | 42 (44%) | 0 | 4 (10.8%) | 18 (66.6%) | 17 (89%) | 3 (60%) | ns |
| Headache, n (%) | 40 (42%) | 1 (14.2%) | 15 (40.5%) | 13 (48%) | 11 (57.8%) | 0 | 0.001** |
| Myalgia, n (%) | 38 (40%) | 2 (28.5%) | 12 (32.4%) | 13 (48%) | 10 (52.6%) | 1 (20%) | 0.003** |
| Arthralgia, n (%) | 36 (37.8%) | 2 (28.5%) | 10 (27%) | 13 (48%) | 10 (52.6%) | 1 (20%) | 0.04* |
| Fatigue, n (%) | 20 (21%) | 0 | 3 (8%) | 8 (29.6%) | 9 (47%) | 0 | ns |
| Rhinorrhea, n (%) | 26 (27.3%) | 1 (14.2%) | 6 (16%) | 12 (44%) | 6 (31.5%) | 1 (20%) | ns |
| Chest pain, n (%) | 13 (13.6%) | 0 | 5 (13%) | 4 (14.8%) | 4 (21%) | 0 | ns |
| Diarrhea, n (%) | 16 (16.8%) | 0 | 4 (10.8%) | 8 (29.6%) | 4 (21%) | 0 | ns |
| Cyanosis, n (%) | 7 (7.3%) | 0 | 0 | 3(11%) | 4 (21%) | 0 | ns |
| Vomiting, n (%) | 5 (5.2%) | 0 | 1 (2.7%) | 3 (11%) | 1 (5.2%) | 0 | ns |
| NA | 105 | 0 | 70 | 12 | 0 | 23 |  |

### The respiratory microbiota composition differs among severity levels for COVID-19 and controls

From the 95 analyzed samples belonging to the upper respiratory tract, we identified a total of 4514 Amplicon Sequence Variants (ASVs). Regarding the analysis of the relative abundance (Fig. 1A), p_Firmicutes, p_Bacteroidetes, and p_Proteobacteria were the most dominant phyla among our severity groups and controls. In general, these phyla are present in all group samples but there are changes in the relative abundance associated with the disease severity. In general, we found p_Firmicutes, p_Actinobacteria, p_TM7, and p_SR1 significantly increased in COVID-19 patients, while p_Bacteroidetes and p_Proteobacteria were found significantly decreased (Supplementary Table S2).

**Figure 1.**
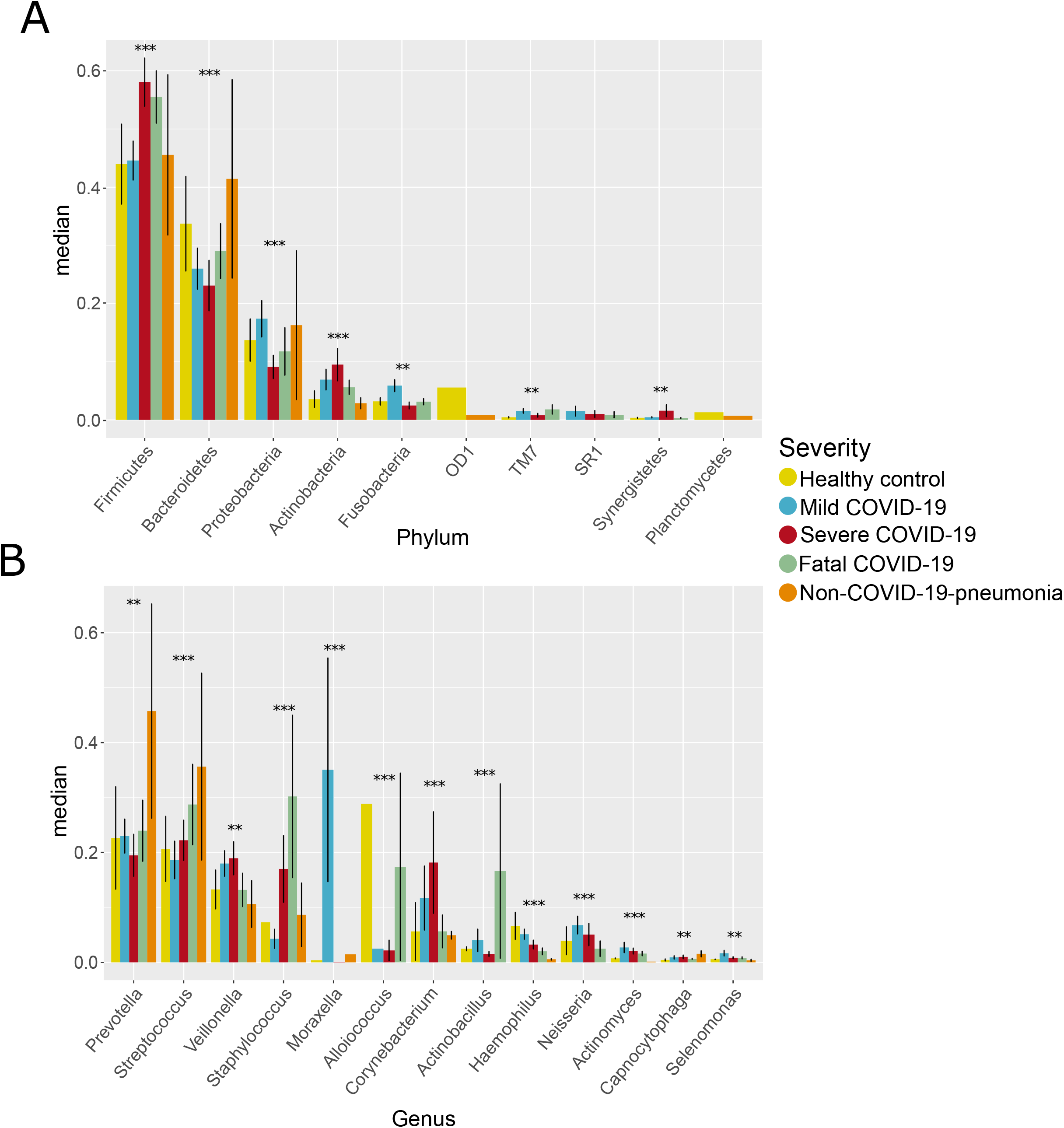
Main composition at phylum and genus level among severity levels for COVID-19 and controls. **A:** median abundance of most abundant phyla in the analyzed groups. **B:** median abundance of most abundant genera in the analyzed groups. Asterisks denote global statistical differences given by Kruskal-Wallis test (*p*-values: * < 0.05, ** < 0.005, *** < 0.0005).

The relative abundance analysis at the genus level revealed genera that significantly differ among COVID-19 patients and controls (Fig. 1B, Supplementary Table S2). In general, we found g_*Veillonella*, g_*Staphylococcus*, g_*Corynebacterium*, g_*Neisseria*, g_*Actinobacillu*s, and g_*Selenomonas* significantly enriched in the COVID-19 patients but reduced in the healthy controls. In contrast, we found g_*Haemophilus* and g_*Alloiococcus* enriched in the healthy controls but reduced in the COVID-19 patients. Moreover, there were differences in the abundance of some genera among the severity levels for COVID-19. For example, g_*Streptococcus* and g_*Staphylococcus* showed an increasing abundance from mild to fatal COVID-19. In contrast, g_*Haemophilus* and g_*Actinomyces* showed the opposite pattern, where the highest abundance is associated with mild COVID-19 and the lowest with the fatal COVID-19. Also, we found g_*Corynebacterium* highly abundant only in severe COVID-19, while g_*Actinobacillus* were found highly abundant only in fatal COVID-19.

Besides, we compared the most abundant phyla for severe and fatal COVID-19 in the upper and lower respiratory tract (Supplementary Figure S3). In particular, we found differences at phylum and genus level. For instance, for severe COVID-19 patients, we found a higher abundance of p_Firmicutes, g_*Neisseria*, and g_*Haemophilus* in the lower respiratory tract. In contrast, we found p_Actinobacteria, p_Fusobacteria, p_SR1, and g_*Staphylococcus* enriched in the upper respiratory tract. For fatal COVID_19 patients, we found p_Proteobacteria, g_*Streptococcus*, g_*Neisseria*, and g_*Capnocytophaga* enriched in the lower respiratory tract albeit, in the upper respiratory tract, we found p_TM7, p_SR1, g_*Corynebacterium*, and g_*Staphylococcus*. Nonetheless, it is worth mentioning that regardless of the differences found in phyla abundance, we found no differences in beta diversity analyses.

#### Alpha diversity

Respecting diversity calculated with the Shannon-Wiener index (Fig. 2), we found healthy controls as the most diverse group and the non-COVID-19-pneumonia group (p < 0.05) as the less diverse. Although among severity groups the differences are not considerable, we did find significant differences among severe and fatal COVID-19 groups (p < 0.05).

**Figure 2.**
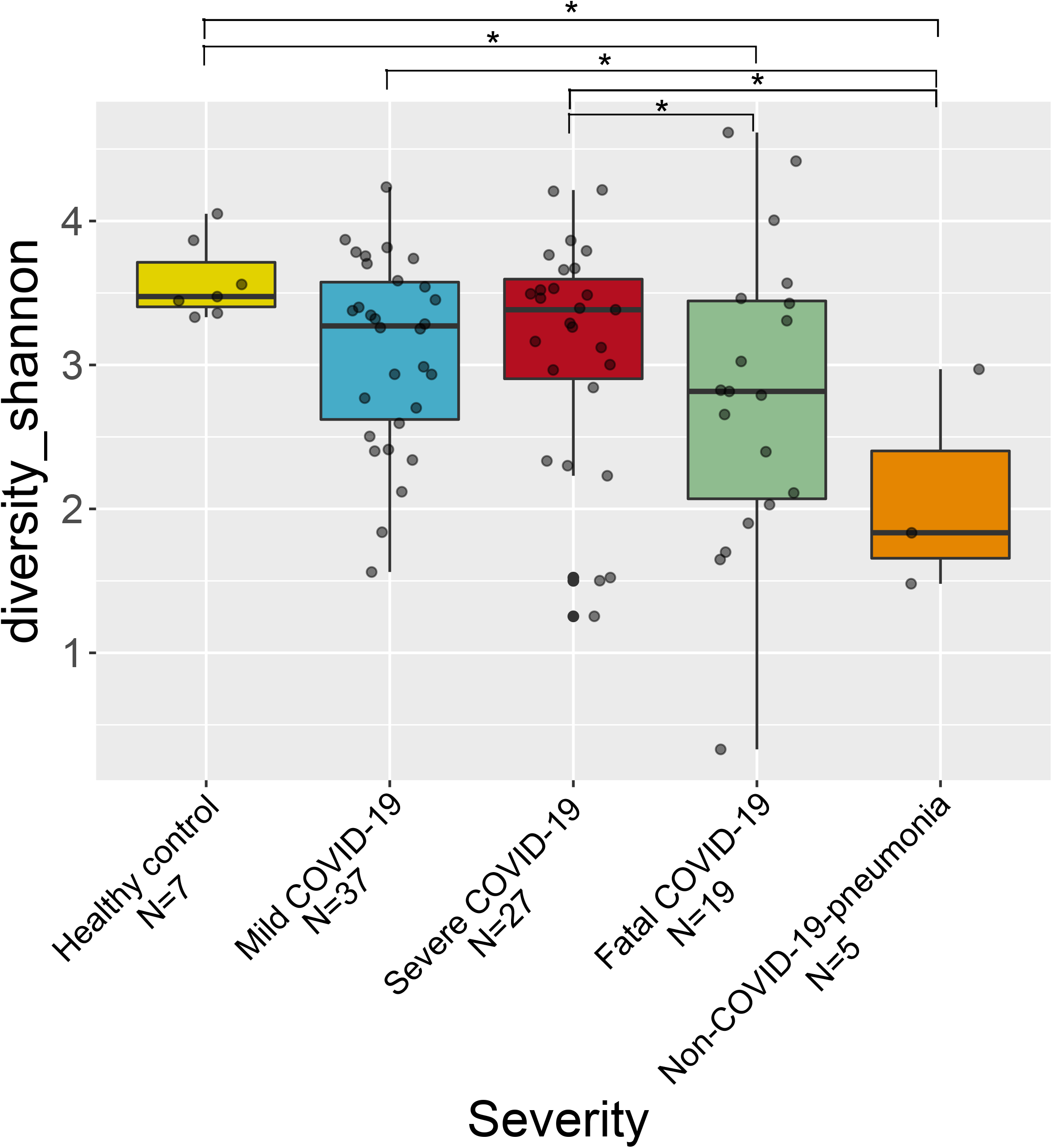
Alpha diversity of the respiratory microbiota among severity levels for COVID-19 and controls. Boxplot of the Shannon-Wiener index value for all analyzed groups. Asterisks denote statistical significant differences given by Wilcoxon rank-sum test (\**p* < 0.05).

#### Beta diversity

The beta diversity analyses showed differences in the microbiota composition among severity levels for COVID-19 and controls (Fig. 3A-B, Supplementary Table S4). Particularly, the PCoA analysis (Fig. 3A) showed differences among severity levels and control groups. Such differences are supported by the PERMANOVA result (F = 2.7, *p* = 0.007). Additionally, the dysbiosis analysis in terms of the Ružička metric allowed us to determine that the microbiota associated with COVID-19 (regardless of severity level) showed significantly higher levels of dysbiosis compared with healthy control (Supplementary Table S4).

**Figure 3.**
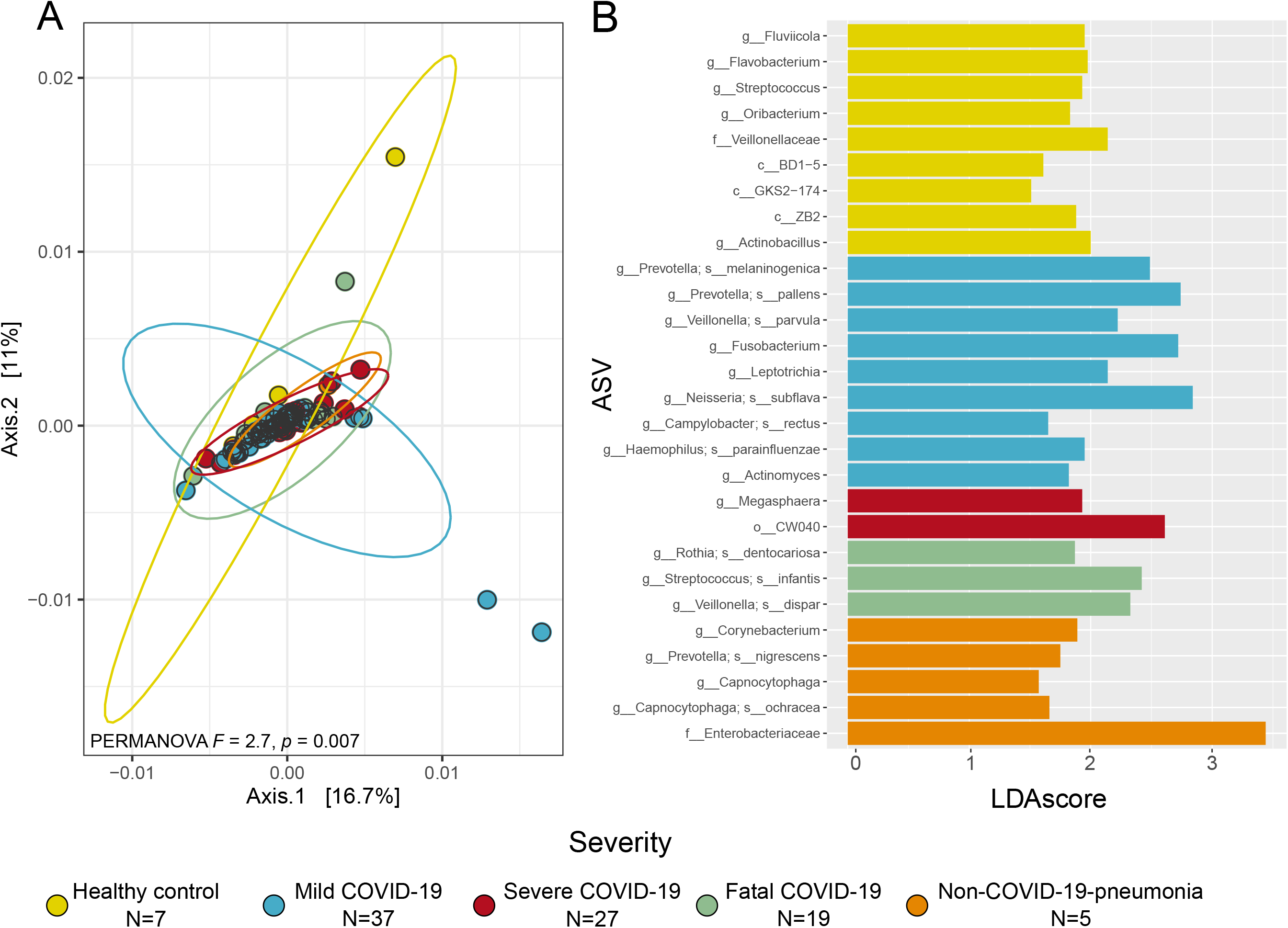
Beta diversity of the respiratory microbiota among severity levels for COVID-19 and controls. **A:** PCoA with weighted Unifrac distance and PERMANOVA result that test differences in the community arrange among groups. Each ellipse represents an analyzed group specified in the legend. **B:** Differentially abundant taxa obtained through LefSe analysis for each group. Only features with a LDA score higher than 1.5 and a *p* < 0.01 were included. ASV: Amplicon Sequence Variant.

The LefSe analysis allowed the identification of differentially abundant taxa associated with the compared groups (Fig. 3B). We observed that all the COVID-19 severity groups and the two control groups showed differentially abundant taxa or biomarkers. In particular, for mild COVID-19, we found g_*Prevotella melaninogenica* and g*_P. pallens*, g_*Veillonella parvula*, g_*Neisseria subflava, g_Fusobacterium, and* g*_Actinomyces* as highly abundant. For severe COVID-19, we found g_*Megasphaera* and o*_*CW040 as the most prevalent. In the case of fatal COVID-19, g_*Rothia dentocariosa*, g_*Streptococcus infantis*, and *g_Veillonella dispar* were the most significant. Moreover, the higher number of differentially abundant taxa were found to be associated with healthy controls (e.g., g_*Streptococcus*, g_*Flavobacterium*, and g_*Oribacterium*, and f_*Veillonellaceae*). Finally, for the non-COVID-19-pneumonia group, we found g_*Corynebacterium*, g*_Prevotella nigrescens*, g_*Capnocytophaga*, and f*_Enterobacteriaceae* as the most abundant.

### Clinical variables associated with mortality risk correlate with specific microbial groups in the respiratory microbiota

The Kaplan-Meier survival curves led to the detection of clinical variables that significantly correlated with survival probability (Fig. 4-A). For instance, we found that APACHE scores above 8 points, levels of Blood Urean Nitrogen (BUN) lower than 40 mg/dl, lymphocytes under 1.25×10^3^/µl, myoglobin above 110ng/ml, troponin above 3.5ng/ml, and urea under 80mg/dl represent high risk by negatively affecting survival probability. The Lefse analysis allowed us to detect bacteria either enriched or depleted in the different risk factor groups for the analyzed variables (Fig. 4-B). We found g_*Neisseria subflava* depleted in the high-risk samples for troponin and APACHE. Moreover, g_*Veillonella dispar* interestingly was found depleted in the low-risk samples for APACHE, BUN, myoglobin, and urea. Nonetheless, we also found some bacterial groups that are constantly enriched in the samples with high-risk for several clinical variables. For instance, g_*Corynebacterium* was found enriched in the high-risk samples for lymphocytes count and urea, while g_*Actinomyces* was enriched in BUN and urea. Besides, four ASV’sof *Prevotella* genus (g_*Prevotella melaninogenica*; g_*Prevotella*; g_[*Prevotella]*; g_[*Prevotella*]_s) were found significantly enriched in the high-risk samples for myoglobin, BUN, troponin, and lymphocytes count.

**Figure 4.**
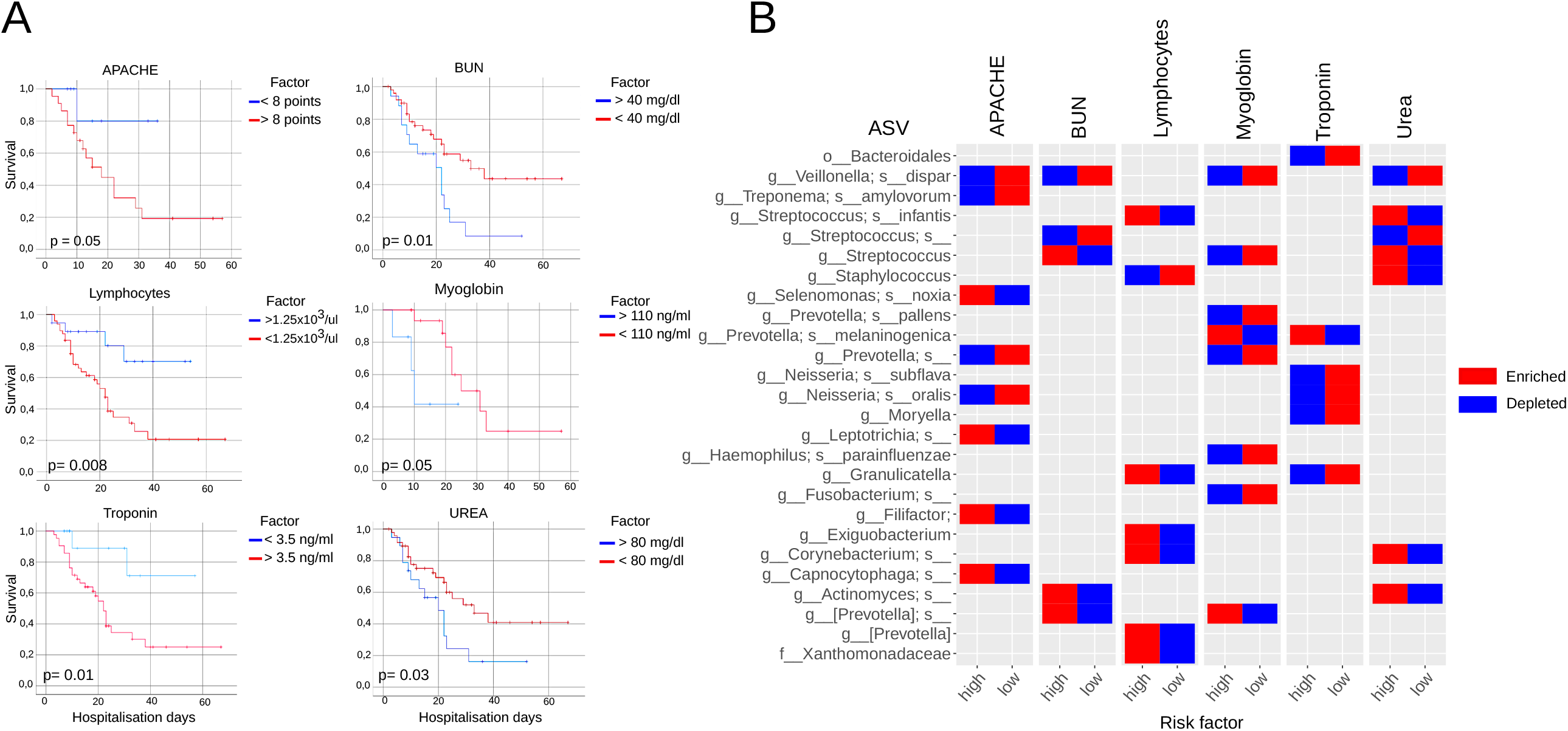
Correlation among clinical variables affecting survival probability and bacteria in the respiratory microbiota. **A:** Kaplan-Meier curves for the clinical variables with a statistical significant difference in survival probability. Variables were classified into two categories specified in the legend “factor”. All curves were constructed with hospitalization days and outcome (either deceased or alive). **B:** Bacteria that are significantly depleted or enriched in samples with the different risk factors for the clinical variables obtained trough Kaplan-Meier curves. Risk factor (either high or low) corresponds to the result of the survival curves (panel A). ASV: Amplicon Sequence Variant. APACHE: Acute Physiology and Chronic Health Evaluation, BUN: Blood Urea Nitrogen.

### The structure of the respiratory microbiota is different among severity levels for COVID-19

The network analysis for the microbiota associated with the severity levels for COVID-19 revealed differences at a structural level (Fig. 5A-B). The graphic representation of the networks showed a different arrangement for each one and a continuum of loss of complexity across COVID-19 severity groups (from mild to fatal) (Fig. 3A). Particularly, the network of the microbiota associated with mild COVID-19 was the largest and more connected one (nodes= 148, edges = 4758) compared to severe COVID-19 (nodes=84, edges=688) and fatal COVID-19 (nodes=74, edges=75). Interestingly, in patients with fatal outcome, the network of the respiratory microbiome was highly disaggregated and poorly connected with multiple isolated nodes (nodes = 74, edges = 75).

**Figure 5.**
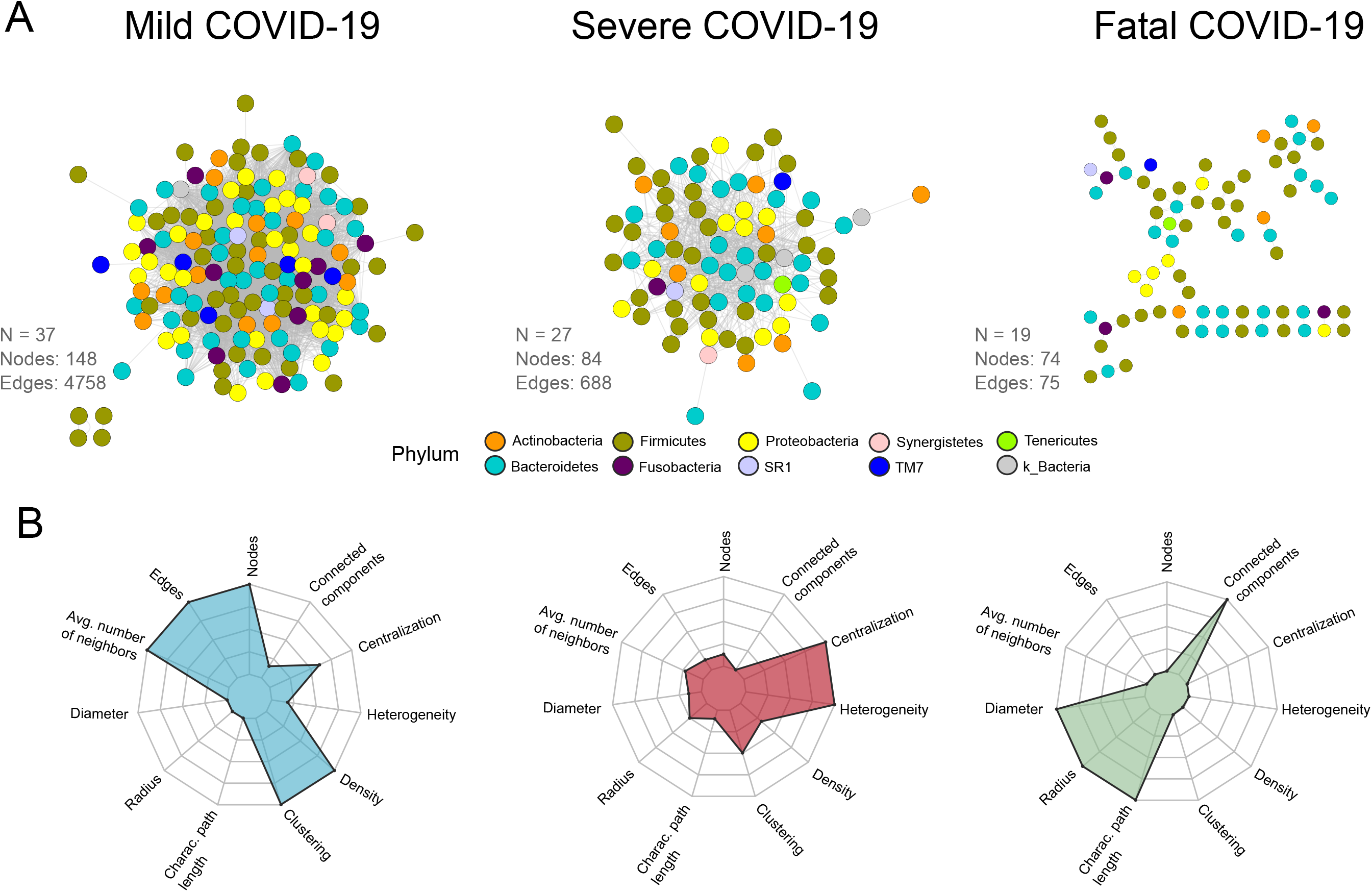
Network structure of the respiratory microbiota among severity levels for COVID-19. **A:** Co-occurrence/exclusion networks for mild, severe and fatal COVID-19. Each node represents a microbial group at ASV level and each edge an interaction (either co-occurrence or co-exclusion). Colors denote phylum identity. Number of samples used to construct the network (N), number of nodes and number of edges are reported in the figure. **B:** Spider chart of the topological metrics associated to each network.

The metric calculation of the networks illustrates that the topology associated with each COVID-19 severity level is different (Figure 3B, Supplementary Table S5). For instance, the mild COVID-19 network exhibited the highest values for the average number of neighbors, density, and clustering. In contrast, the severe disease network was characterized by greater centralization, and heterogeneity, while the fatal disease network showed the highest values for diameter, characteristic path length, and connected components.

## Discussion

COVID-19 pandemic has raised major scientific efforts to identify factors associated with the different disease severity outcomes. Here, we characterized the respiratory microbiota as a potential factor affecting severity outcome for COVID-19. We assessed differences in diversity and structure of the microbial communities associated with a large cohort of patients and linked the results to clinical variables to get insights into the mechanisms by which the microbiota may impact host response against the disease.

As well in recent studies investigating COVID-19 etiology ^3^, we found that demographic and health-related factors showed strong associations with severity. Male sex, high values for BMI, age above 50 years old, and previous antibiotic treatment were significantly associated with fatal COVID-19 patients (Table 1), thus potentially favoring the development of a fatal state of the disease.

Likewise in previous work exploring COVID-19 associated respiratory microbiota ^14,15^, we found significantly lower microbial diversity in the microbiota of COVID-19 patients than in the healthy controls (Fig.1A). This result is relevant since, in general, more diversity is correlated with a better response of the microbial systems against perturbance (e.g., disease). A more diverse microbiota can persist after disease (e.g., by functional redundancy) or recovers to an earlier state (e.g., resilience) ^16^ having direct consequences on the host’shealth ^8^.

Furthermore, we found differences in the abundance of some bacteria among our study groups (Fig. 1A, Supplementary Table S2). In particular, as well as other respiratory diseases ^17^, we observed an increased ratio of p_Firmicutes/p_Bacteroidetes in COVID-19 patients. p_Firmicutes was detected highly abundant while p_Bacteroidetes where particular decreased in the microbiota associated to COVID-19 patients. This is of particular interest since, in murine models, it has been proven that p_Bacteroidetes can down-regulate the expression of ACE2 ^18^. Although the correlation was observed in gut microbiota, the particular low abundance of members of such phylum in severe and fatal patients in this work opens the question about if this process could also take place in the respiratory tract.

Regarding the analysis at the genus level, the most significant differences were found in potentially pathogenic bacteria (Fig. 1B, Supplementary Table S2). We identified a gradual increase of g_*Streptococcus* from mild to fatal COVID-19. Although g_*Streptococcus* is usually a commensal member of the respiratory microbiota, it can become pathogenic in the face of environmental disturbs. In higher abundance, such genus it has been linked to viral acute respiratory infections ^19,20^. Furthermore, genera such as g_*Veillonella*, g_*Staphylococcus*, and g_*Actinomyces* also exhibited high abundance in the different severity levels for COVID-19. Specifically, g_*Veillonella* and g_*Actinomyces* have been found as opportunistic pathogens in COVID-19 ^21^. Moreover, g_*Staphylococcus* is one of the most common causal agents of secondary infections in respiratory diseases such as influenza ^22^.

Regarding beta diversity, we found some differences among the analyzed groups. For instance, we observed in the PCoA analysis that the samples belonging to severe and fatal COVID-19, as well as to the non-COVID-19-pneumonia group, are more alike in terms of microbial composition than the healthy controls and the mild COVID-19 patients (Fig. 2A). Moreover, our dysbiosis analysis let us detect that the microbiota of COVID-19 showed higher levels of dysbiosis than the healthy control (Supplementary Table S4). Several MADs exhibit this behavior in which microbiota instability (dysbiosis) is present not by the dominance of one or a few bacteria but by a higher heterogeneity/stochasticity of microbial groups ^9^.

Dysbiosis implies a disruption in the bidirectional interactions between the host immune system and the microbial communities, potentially altering functions provided by these communities and reshaping the whole host-microbiota interaction ^8,23^. It has been shown that microbiota stability is a hallmark of the health and homeostasis of the host ^6,8,20^. For instance, some reports suggest that there is a homeostatic mechanism that keeps lung epithelium in an interferon prime state with antiviral activity against other respiratory infections such as influenza. This particular antiviral response is stimulated by specific pathogen-associated molecular patterns (PAMPs) that are induced by the microbial communities ^24^. In this sense, the modification of microbial communities (e.g., dysbiosis) showed in COVID-19 patients could be modifying the specific PAMPs that stimulate this homeostatic antiviral response, allowing for better conditions for respiratory virus infection, encompassing SAR-CoV-2.

A common question when studying MADs is whether dysbiosis enhances disease or is caused by it. For COVID-19, the clinical outcome is highly correlated with comorbidities such as hypertension, diabetes, and obesity ^7^, which are often associated with dysbiosis in the gut microbiota ^21^. This remark, together with the highly distributed antibiotic uptake in COVID-19 patients (53.6% in our cohort, regardless of severity), merits a reflection on the possibility that most of the patients could be dysbiotic at the time of the disease. In other respiratory diseases such as COPD and asthma, it has been shown that dysbiosis in the respiratory microbiota can lead to a deregulated immune response, increasing inflammatory processes ^6,8,25^. Considering that aberrant immune responses are determinant in COVID-19 progression, a previous dysbiotic respiratory microbiota could be affecting disease progression.

The LefSe analysis (Fig. 3B) shows a differential abundance of microbial groups. For example, we found that most of the groups associated with the healthy controls belong to the so-called “normal” respiratory microbiota (e.g., g_*Streptococcus, g_Oribacterium*, and f_*Veillonellaceae*) ^26^. In contrast, when we look at the results of the microbiota associated with COVID-19 and non-COVID-19-pneumonia groups, other potentially pathogenic microbial groups appear. In particular, in patients with non-COVID-19-pneumonia we found bacteria associated with nosocomial infections such as g_*Corynebacterium* ^27,28^. For mild COVID-19, we found some microbial groups associated with disease or bacteremia like g_*Prevotella melaninogenica*, g_*V. parvula* and g_*Neisseria subflava* ^7,28^. For the case of severe COVID-19, we found g_*Megasphaera* that has been associated with the risk of ventilator-associated pneumonia (VAP) in other studies characterizing COVID-19 microbiota ^12^. Additionally, we found g_*Rothia dentocariosa* highly abundant in deceased patients. This bacteria has been found as the causal agent of secondary pneumonia in H1N1 infection ^22^ and more recently have been associated with disease progression in previous studies characterizing COVID-19 respiratory microbiota, being proposed as a biomarker for the disease ^13,14^.

From a clinical standpoint, it makes sense that a higher mortality predictor such as APACHE score correlates with low survival in COVID-19 patients. Other clinical factors such as BUN or urea have also been used as severity markers in respiratory diseases such as community-acquired pneumonia ^29^. Acknowledging that multiple pathophysiological considerations still unexplained in SARS-CoV-2 infection, and the multisystemic involvement that has been observed in COVID-19 ^30^, biochemical markers of organ dysfunction such as lymphopenia, elevated myoglobin, and troponin serum levels as those found in this study, can help predict mortality in these patients ^31^. Although the association of these factors with specific microbial groups in the respiratory tract has not been previously reported, the findings in this study open the path to further study the relationship between respiratory microbiota and clinical outcomes. The identification of pathogenic bacteria such as g_*Actinomyces* g_*Prevotella* and g_*Corynebacteriu*m in association with two or more clinical factors further supports the current research line trying to correlate the gut-lung axis with pulmonary disease ^32^.

Recent studies, as well as this work, suggest that particularly anaerobic bacteria inhabiting the respiratory tract may be involved in COVID-19 pathogenesis and host immune system. In particular, *g_Prevotella* has been found to increase in studies with patients with severe disease and have been co-related to cardiac injury and higher risk mortality ^31,33^. In this work, we found this specific genus associated with four clinical variables that predict mortality in patients with COVID-19 (Fig. 4A). This finding is of special interest considering previous evidence of g_*Prevotella* enhancing a Th17 mediated response through IL-8, CCL20, and IL-6 secretion ^33,34^; both the Th17 response and its cytokines are currently associated with the host’s immune response to SARS-CoV-2 ^35^.

Finally, the co-occurrence arrangement of ecological networks lets us identify structural patterns that reflect variations in the biological properties of the microbial communities associated with COVID-19. For instance, we found that all networks are distinguishable in terms of topological metrics such as density, clustering, and heterogeneity. It is worth mentioning that such metrics are potentially related to the stability of the systems likewise to other ecological properties such as resilience and redundancy ^36^. In particular, we found a striking pattern of reduction of structural complexity from mild to fatal COVID-19. The loss of complexity is showed by a reduction in the number of nodes, edges (connections), density, and clustering. Passing through from a highly connected and dense network (mild COVID-19) to a highly disaggregated, unconnected network (fatal COVID-19) (Fig. 5AB, Supplementary Table S5).

Those structural changes can lead to the generation of hypotheses regarding consequences at the microbial community level. For instance, changes in structural patterns could potentially be reflected in alterations in the ecological relationships among microorganisms. A common feature in MADs is that commensal/neutral bacteria can become pathogenic at the face of disease ^21^. That is the case of bacteria such as g_*Prevotella*, g_*Veillonella*, g_*Streptococcus*, g*_Actinomyces*, or g_*Megasphaera*, which have been found as opportunistic pathogens in other studies characterizing COVID-19 microbiota ^12,19–21^ and that were also found in this work (severe and fatal associated microbiota (Fig. 2B)). The shift from neutral to deleterious interactions in specific bacteria could be the result of a loss of interactions that maintain the function and stability of the microbial systems. Which in turn, could cause an exacerbate growth of microbial groups potentially pathogenic, but also the depletion of beneficial bacteria, altering the whole environment and possibly compromising functions provided by the microbiota to the host.

## Conclusions

Overall, this work provides insights into the role of the respiratory microbiota in COVID-19 disease. Our data suggest that host-related and environmental factors could be affecting the respiratory microbiota before SARS-CoV-2 infection, potentially compromising the immunological response of the host against disease and promoting secondary bacterial infections. For instance, the high levels of dysbiosis coupled with poor microbial structural complexity in the respiratory microbiota of COVID-19 patients, possibly resulted from antibiotic uptake and comorbidities, could have consequences at the host and microbial community level. On the one hand, increased dysbiosis in diseased patients could be modifying the PAMPs that stimulate a homeostatic antiviral response, allowing for better conditions for SAR-CoV-2 replication. Additionally, the loss of structural complexity may provoke the appearance of opportunistic pathogens that, through ecological competition, can cause the depletion of beneficial bacteria and promote secondary bacterial infections that worsen the clinical outcome. In summary, the findings of this work contribute to understand the pathology of COVID-19 by identifying the respiratory microbiota as a potential factor affecting disease outcome. Further investigations looking for the specific mechanisms by which dysbiotic microbiota in the respiratory tract compromise immunological responses against virus infections are needed.

## Methods

### Ethics statement

The Science, Biosecurity and Bioethics Committee of the Instituto Nacional de Enfermedades Respiratorias revised and approved the protocol and the consent procedure given by the participants or their legal guardians (B-0520). Additionally, the Institution requested an informed consent for the recovery, storage, and use of the biological remnant to research purposes.

### Study design

As part of a surveillance program at the Instituto Nacional de Enfermedades Respiratorias Ismael Cosío Villegas (INER), 115 initial respiratory samples (oropharyngeal swabs, nasopharyngeal swabs, and tracheal aspirates) were collected between March 2020 and October 2020. Additionally, we included seven subjects without respiratory symptoms and negative SARS-CoV-2 RT-PCR test (healthy), and five patients with pneumonia that were hospitalized but negative to SARS-CoV-2 (non-COVID-19-pneumonia control group). Patients with COVID-19 were classified into three mutually exclusive categories of severity: a) mild COVID-19 (patients with moderate symptoms that did not required hospitalization), b) severe COVID-19 (patients that required hospitalization and were subject to Invasive Mechanical Ventilation (IMV)), and c) fatal COVID-19 (deceased patients). Overall, a total of 37 patients with mild disease, 38 with severe disease and, 40 with fatal outcome were included in the study.

### DNA extraction and 16S rRNA sequencing

Respiratory samples, either nasopharyngeal swabs, oropharyngeal swabs or tracheal aspirates, for all 127 patients were collected and centrifugated for 15 min at 4,800 g, and the pellet was used for DNA extraction. DNA was extracted using the QIAmp Cador Pathogen Mini Kit extraction (Qiagen N.V., Hilden, Germany) according to the manufacturer’sinstructions. V3-V4 16S rRNA region was amplified by PCR using the primers reported by Klindworth et al (2013) ^37^ (for more information see Supplementary Material S1). Library preparation was done according to the Illumina 16S metagenomic sequencing protocol with few modifications. Briefly, 16S amplicons were purified with the DNA clean & concentrator kit (Zymo Research, Irvine Cal., USA). Dual indices and Illumina sequencing adapters were attached in a second PCR step using Nextera XT Index Kit V2 (Illumina, San Diego Cal., USA). Finally, amplicons were purified, pooled in equimolar concentrations, and sequenced in a MiSeq Illumina instrument generating paired-end reads of 250bp.

### Sequence data processing

Illumina raw sequences were processed with QIIME2 (v.2020.8) ^38^. Sequences denoising, quality filtering, and chimera checking were performed with DADA2 ^39^. From the original number of reads (13,533,440), we kept a total of 9,499,204 with an average of 73,637 sequences per sample. The Amplicon Sequence Variants (ASVs) were aligned with MAFFT ^40^ and used to construct a phylogeny with fasttree2 ^41^. ASVs taxonomy was assigned with the Näive Bayes classifier *sklearn* ^42^ using the Greengenes 13.8 database ^43^. All ASVs identified as mitochondria (N=10), and chloroplast (N=32) were removed. Raw data were deposited in the NCBI Sequence Read Archive (SRA) (PRNAJ726205).

### Diversity, compositional, and statistical analyses

Since our sample set contains both upper (Oropharyngeal swabs [OPS], Nasopharyngeal swabs [NPS]) and lower (Tracheal aspirates [TA]) respiratory tract samples, and it is known that these sites vary in microbial composition, we used only upper respiratory samples for the main analyses. After this process we kept a total of 95 samples (mild COVID-19 = 37, severe COVID-19 = 27, fatal COVID-19 = 19, healthy control = 7, and non-COVID-19-pneumonia = 5). We also characterized TA samples to compare levels of severity in the microbiota of the upper and lower respiratory tract.

#### Composition analyses

In order to determine if the samples associated with different severity levels and controls differed in the most abundant phylum and taxa, we plotted a bar graph by using the median and standard error of each taxon in the analyzed groups. Besides, we performed a Kruskal-Wallis test to determine if there were any significant differences followed by a paired Wilcoxon rank-sum test in the “vegan” R package ^44^.

#### Alpha diversity

We calculated the Shannon-Wiener diversity index with the “microbiome” R package ^45^. To detect potential differences among groups we conducted a Wilcoxon rank-sum test in the “vegan” R package ^44^.

#### Beta diversity

We carried out a Principal Coordinates Analysis (PCoA) with weighted UniFrac distance at ASV level in the “phyloseq” R package ^46^. Potential differences in beta diversity were addressed with a Permutational Analysis of Variance (PERMANOVA) with 999 permutations performed with the “vegan” R package ^44^. Additionally, we tested for dispersion and stochasticity as a proxy of dysbiosis in microbial communities ^47^. For this, we calculated the Ružička similarity metric in the “CommEcol” R package ^48^ and performed a Wilcoxon rank-sum test to detect potential statistical differences between healthy controls and diseased groups in their intra-treatment sample similarities. Dysbiosis was assumed when the similarities between the healthy microbiota samples were significantly higher than the similarities between the diseased microbiota samples ^9^.

Finally, to detect differentially abundant taxa associated with severity levels and controls, we performed a Linear Discriminant Analysis (LDA) with effect size (LefSe) at the ASV level using the web-based tool MicrobiomeAnalyst ^49^. Only taxa with a LDA score higher than 1.5 and a *p*-value < 0.01 were used. All diversity and statistical analyses were performed in *R* program (v.1.3.1) ^50^.

### Clinical data analyses

In order to analyze the clinical data associated with our patient’scohort, we transform each clinical variable into a binomial category according to its data distribution. We used cut-points based on the 25 and 75 percentiles for each variable. For example, for a given variable, we classified all samples with values above or equal to the 75 percentile as “1”, and all samples with values under the 75 percentile as “2”. Subsequently, we constructed Kaplan-Meier survival curves in SPSS Statistics (version 21) (Chicago, Illinois, USA) by using the hospitalization days as time variable, the mortality status (either deceased or alive) as a dependent variable, and the specific clinical qualitative variables as exposure variable. Only those curves statistically (*p* < 0.05) and biologically meaningful were retained for subsequent analyses.

Also, to determine if there were differentially abundant bacteria associated with the several risk factors for the clinical variables obtained from the Kaplan-Meier curves, we performed a second LefSe analysis. From this result, only taxa with a LDA score higher than 1.5 and a *p*-value < 0.01 were used.

### Network structure inference

We inferred the network structure for the microbiota associated with the different severity levels. Network calculation was performed in the software CoNet^36^ by using read counts summarized at the ASV level. One network was constructed for each severity level (samples; mild COVID-19: 37, severe COVID-19: 27, and fatal COVID-19: 19). Only co-occurrences statistically supported by the three tested methods (Pearson, Spearman, and Kendall) with a correlation > 0.85 and a *p*-value < 0.01 were established as edges in the graphs. Also, we applied a multi-test correction using the Benjamini-Hochberg procedure. Network visualization was performed in Cytoscape (v. 3.8.2) ^51^.

To further characterize the structure, we computed metrics of the topology of each network using the NetworkAnalyzer plug-in in Cytoscape ^51^ and visualized them with a spider chart constructed in R program.

## Data Availability

The sequencing reads generated during the present study are available in the NCBI Sequence Read Archive (SRA) accession PRNAJ726205.

## Acknowledgments

We are thankful for the excellent clinical care of all physician and nurses at INER attending COVID 19 patients.

## Author contributions

AHT, JSH, RPP, and JAVP conceived and designed the project. OB, EG, EBV, CGP, and VHAT collected the clinical data and constructed the database. AHT, FMN, CB, MM, ARR, and JVP performed the experimental laboratory procedures. AHT and MC performed the bioinformatics and statistical analyses. MTH, RS, SR, and JAVP performed the interpretation of clinical data. AHT, FMN, MTH, SAR, RS, SR, RPP, and JAVP wrote the manuscript. JAVP supervised the project and led the team. All authors discussed the results and commented on the manuscript.

