## Supplementary Material for "Dysbiosis and structural disruption of the respiratory microbiota in COVID-19 patients with severe and fatal outcomes"

#### S1. Specificis for 16S rRNA gene amplification

**PCR cycles:**

95°C for 3 min, followed by 35 cycles at 95° C for 30s, 55°C for 30s, and 72°C for 30s, with a final extension at 72°C for 5 min

**Primers used:**

F - 5´ CCTACGGGNGGCWGCAG 3´

R - 5´GACTACHVGGGTATCTAATCC 3´

### Table S2. Wilcoxon paired comparisons of taxa abundance among groups.

| Taxa/ Comparison | Healthy vs Mild | Healthy vs Severe | Healthy vs Fatal | Healthy vs Non-COVID-19-pneu. | Mild vs Severe | Mild vs Fatal | Mild vs Non-COVID-19-pneu | Severe vs Fatal | Severe vs Non-COVID-19-pneu. | Fatal vs Non-COVID-19-pneu |
|---|---|---|---|---|---|---|---|---|---|---|
| **Phylum** | | | | | | | | | | |
| Firmicutes | NS | NS | 0.03* | 0.04* | 0.05* | NS | NS | 0.02* | 0.03* | NS |
| Bacteroidetes | 0.03* | NS | 0.02* | 0.02* | NS | NS | NS | NS | NS | NS |
| Proteobacteria | 0.01* | NS | 0.04* | 0.03* | 0.001** | NS | NS | NS | NS | NS |
| Actinobacteria | NA | NA | NA | NA | NS | NS | 0.003** | NS | 0.007** | 0.004** |
| Fusobacteria | 0.004** | 0.02* | NS | 0.0003*** | NS | 0.02* | 0.0004*** | NS | 0.0001*** | 0.0004*** |
| TM7 | 0.0001*** | 0.002** | 0.006** | NA | 0.03* | 0.01* | NA | 0.008** | NA | NA |
| Synesgistetes | NS | 0.01* | NS | NA | 0.004** | NS | NA | 0.002** | NA | NA |
| OD1 | NA | NA | NA | 0.0001*** | NA | NA | NA | NA | NA | NA |
| GN02 | NA | NA | NA | 0.0003*** | NA | NA | NA | NA | NA | NA |
| Plantomycetes | NA | NA | NA | 0.002** | NA | NA | NA | NA | NA | NA |
| **Genus** | | | | | | | | | | |
| *Prevotella* | NS | NS | NS | 0.002** | NS | NS | 0.001** | NS | 0.002** | 0.003** |
| *Streptococcus* | NS | 0.006** | NS | NS | 0.01* | NS | 0.04* | 0.02* | 0.01* | NS |
| *Veillonella* | 0.002** | 0.003** | 0.006** | 0.002** | NS | NS | 0.004** | NS | 0.003** | 0.008** |
| *Staphylococcus* | 0.0004*** | 0.003** | 0.0008*** | 0.005** | 0.0007*** | 0.0001*** | 0.006** | 0.002** | NS | 0.01* |
| *Moraxella* | 1.2e-05*** | NS | NA | 0.003** | 1.8e-07*** | NA | 2.3e-04*** | NA | 0.0003*** | NA |
| *Alloiococcus* | 3.2e-09*** | 2e-08*** | 1.6e-05*** | NA | NS | 0.0005*** | NA | 0.003** | NA | NA |
| *Corynebacterium* | 0.0002*** | 1.7e-04*** | 0.02** | 0.008** | 0.03* | 0.02* | 0.02* | 0.001** | 0.002** | NS |
| *Actinobacillus* | NS | 0.02** | 0.001** | NA | 0.03* | NS | NA | 0.05* | NA | NA |
| *Haemophilus* | NS | NS | 0.02* | 0.001** | NS | 0.05* | 0.03* | NS | 0.001** | 0.003** |
| *Neisseria* | 0.01* | 0.01* | NS | NA | NS | NS | NA | NS | NA | NA |
| *Magasphaera* | NS | NS | NS | 0.001** | NS | NS | 0.002** | NS | 0.004** | 0.001** |
| *Actinomyces* | 0.008** | 0.006** | 0.009** | 0.02* | NS | NS | 0.03* | NS | 0.03* | 0.007** |
| *Capnocytophaga* | NS | NS | NS | NS | NS | NS | NS | NS | NS | NS |
| *Selenomonas* | 0.003** | 0.02* | 0.02* | NS | NS | NS | 0.01* | NS | 0.05* | NS |

*p*-values corresponds to paired Wilcoxon rank-sum test.
NS= non-significant *p*-value
NA= comparison not available due to absence of such taxa in one of the comparison group.

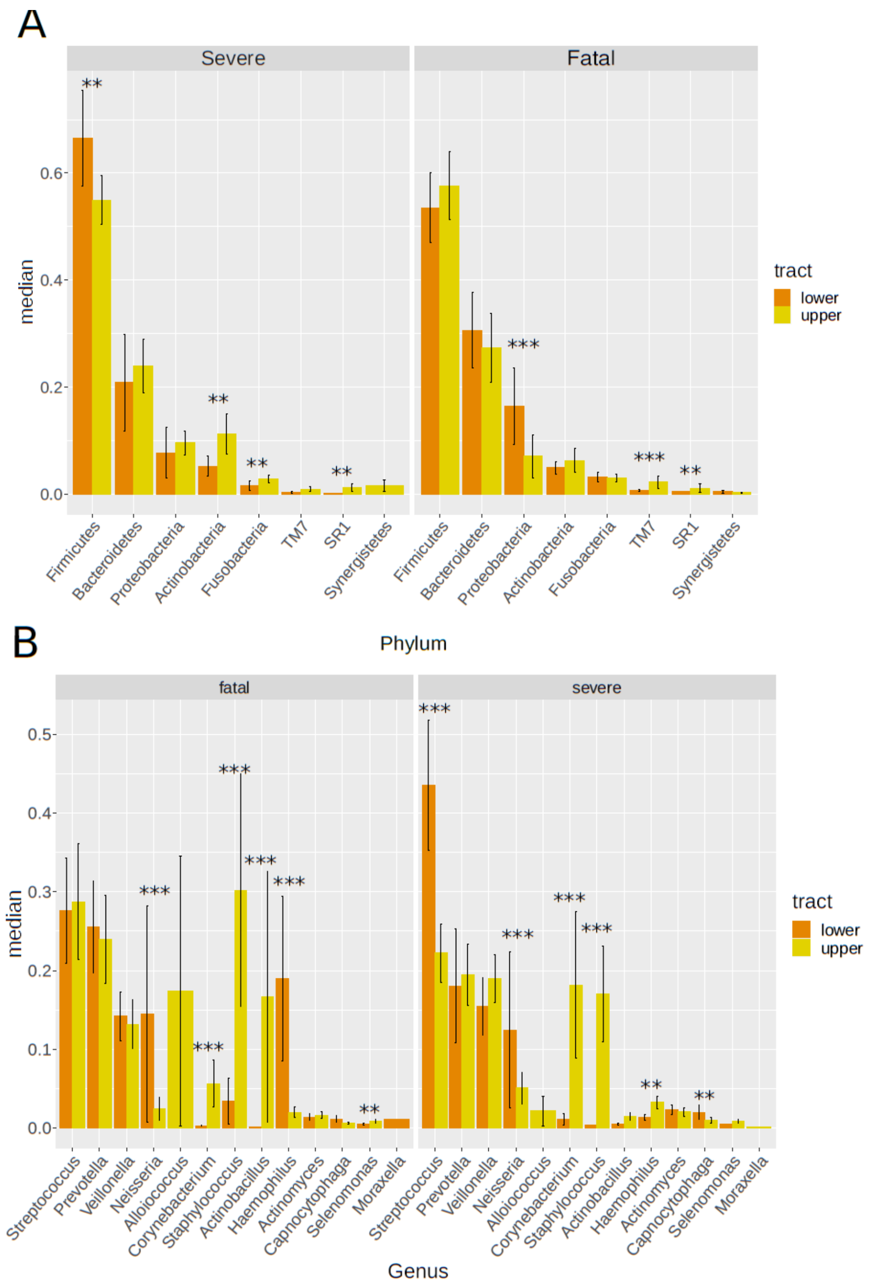


**Figure S3. Main composition at phylum and genus level between lower and upper respiratory tract. A:** median abundance of most abundant phyla in severe and fatal COVID-19. **B:** median abundance of most abundant genera in severe and fatal COVID-19. Asterisks denote statistical differences given by Wilcoxon rank-sum test test (*p*-values, * < 0.05, ** < 0.005, *** < 0.0005).

**Table S4. Ružička metric as a proxy of dysbiosis among severity levels for COVID-19 and controls.**

| Comparison | | Mean similarity | | |
|---|---|---|---|---|
| **Healthy** | **Diseased** | **Healthy** | **Diseased** | **Wilcoxon Test *p* value** |
| Healthy control | Mild COVID-19 | 0.94 | 0.89 | 0.05* |
| | Severe COVID-19 | 0.94 | 0.88 | 0.01* |
| | Fatal COVID-19 | 0.94 | 0.89 | 0.05* |
| | Non-COVID-19-pneumonia | 0.94 | 0.97 | 0.08 |

Intra-treatment similarity for healthy and diseased microbiota in terms of Ružička distance. Statistically significant differences were accessed with Wilcoxon rank-sum test. Dysbiosis was assumed when the similarities between the healthy microbiota samples were significantly higher than the similarities between the diseased microbiota samples (more dispersed).

**Table S5. Topological metrics for all the calculated co-occurrence networks.**

| Metric | Mild COVID-19 | Severe COVID-19 | Fatal COVID-19 |
|---|---|---|---|
| Number of nodes | 148 | 84 | 74 |
| Number of edges | 4758 | 688 | 75 |
| Average number of neighbors | 65.86 | 16.35 | 2.31 |
| Diameter | 4 | 5 | 11 |
| Radius | 2 | 3 | 6 |
| Characteristic path length | 1.59 | 1.94 | 4.99 |
| Clustering | 0.788 | 0.47 | 0.155 |
| Density | 0.461 | 0.197 | 0.075 |
| Heterogeneity | 0.574 | 0.856 | 0.512 |
| Centralization | 0.391 | 0.563 | 0.127 |
| Connected components | 3 | 1 | 14 |
